# The Effect of Cryo-auriculotherapy on Post-Operative Pain Management following Rotator Cuff Surgery: A Randomized, Placebo-Controlled Study

**DOI:** 10.1101/2022.02.05.22270330

**Authors:** Jacques E. Chelly, Steven L. Orebaugh, Mark W. Rodosky, Yram J. Groff, Brittany E. Norton, Amy L. Monroe, David Alimi, Senthilkumar K. Sadhasivam, Keith M. Vogt

## Abstract

**Background:** In the context of the current opioid crisis, there is a growing interest in evaluating non-pharmacological solutions to manage post-operative pain. Evidence supports the concept that auriculotherapy may provide significant analgesia

**Objective:** Investigating the efficacy of cryo-auriculotherapy to reduce postoperative opioid consumption following a rotator cuff surgery.

**Settings:** Academic medical center, USA

**Methods:** This trial was registered to Clinicaltrials.gov (NCT03860259). A total of 39 subjects undergoing rotator cuff surgery and randomized to receive either an active cryo-auriculotherapy treatment (Auriculotherapy; n=20) or a placebo treatment (placebo; n=19) were included in the analysis. For each cryo-auriculotherapy subject, the treatment was performed in the recovery room. The primary endpoint was overall opioid consumption (oral morphine equivalent = OME). Secondary endpoints included pain and overall non-narcotic analgesic consumption on postoperative day 5, patient satisfaction and function recovery using the 12-Item Short Form Health Survey (SF-12), time to discharge from the recovery room and the hospital and patient satisfaction, as well as the number of subjects from each group readmitted because of pain-related issues.

**Results:** The use of cryo-auriculotherapy was associated with a 35% decrease in total opioid requirement over the first five-day recovery period and a 15% decrease in pain with movement. Pain with movement in the auriculotherapy group remained lower compared to the placebo group for at least 14 days (4.47 ± 2.12 vs 5.84 ± 2.39, respectively; p=0.0394).

**Conclusions:** Our data suggests that cryo-auriculotherapy represents an alternative to opioids in patients undergoing rotator-cuff surgery.

## INTRODUCTION

The current opioid epidemic has led to a renewed interest in exploring non-pharmacological techniques to treat post-operative pain. ^(1)^ Furthermore, the use of opioids is associated with significant adverse effects in the context of surgery, including postoperative nausea and vomiting, respiratory depression, hypotension, immunosuppression, and constipation. ^(2-5)^ Although interscalene block is the gold standard for postoperative pain management following shoulder surgery, the duration of the block only covers the initial rehabilitation period, and in most cases, patients require the use of opioids.^(6)^

Auriculotherapy is a non-invasive, complementary technique that has been studied in several surgical models, including tooth extraction, ^(7,8)^ hip arthroplasty, ^(9)^ gynecologic procedures, ^(10, 11)^ and depression. ^(7, 12)^ All recent reviews have concluded that additional evidence is required to determine the role that such a technique may play in perioperative pain management because there is still a limited number of randomized, placebo-controlled studies ^(13-16)^ on its use. Furthermore, several techniques are reported to interact with the ear points included in the proposed treatment,including needles^(17)^,, lasers^(18)^, magnetic sticking ^(10)^, and electrostimulation^(19)^.

The current study was designed to assess the role that auriculotherapy may play in reducing post-operative pain and opioid requirement in opioid-naïve patients undergoing elective primary rotator cuff repair with our standard protocol that includes interscalene block and general anesthesia.

## METHODS

This was a prospective, randomized, placebo-controlled trial conducted at the University of Pittsburgh Medical Center (UPMC) Shadyside and Montefiore hospitals. The protocol was reviewed and approved by the Institutional Review Board (IRB) of the University of Pittsburgh Human Resources Protection Office (STUDY18050099) and was registered to Clinicaltrials.gov (NCT03860259) before any eligible patients were approached and consented.

### Recruitment

Patients were recruited in the pre-operative holding area of UPMC Shadyside or UPMC Montefiore on the day of their scheduled rotator cuff surgery. Patients who agreed to participate signed an IRB-approved informed consent form and were randomized to receive either an active auriculotherapy treatment (auriculotherapy group) or a placebo auriculotherapy treatment (placebo group) in the recovery room. The randomization sequence was determined using computer-generated random numbers. Each patient was made aware at the time of consent and throughout the study that they could choose to withdraw at any time.

### Inclusion Criteria

Patients over 18 years of age undergoing elective rotator cuff repair and consented for an interscalene block performed prior to the transfer to the operating room were eligible for study inclusion.

### Exclusion Criteria

Chronic pain requiring daily opioid use; history of opioid use disorder; contraindications for interscalene block, including allergy to local anesthetics; anatomical malformation that the investigators felt may interfere with performing the nerve block; Raynaud’s disease diagnosis, and peripheral vascular disease.

### Treatment Groups

Auriculotherapy consisted of the treatment of nine ear points. The location of these points was based on cartography developed by Alimi D. ^(20)^ These points included Ω2 (the master point for the mesoderm; A4), the shoulder sensory point (B11), six points involved with the pain pathway at the level of the brainstem and the spine (stellar ganglion (F10); C7, sensory (C11) and motor C7, (CXI), sensory master point (SMP; D17), the reticular master point (RMP, H13), thalamus (G14)), and ACTH (I17).

The auriculotherapy treatment was performed in the recovery room on the ear ipsilateral with the surgery site using a cryoauriculopunctor (Matauris, Maison Alfort, France). The cryoauriculopunctor was comprised of an injector connected to a nitrogen gas canister. Each subject was randomized to receive either an active treatment (auriculotherapy group) or an inactive treatment (placebo group). The active auriculotherapy treatment consisted of delivering a 2 s jet of nitrogen gas at the level of each ear point, whereas the placebo treatment consisted of delivering a 2 s jet using an empty canister at the level of each point. The cryoauriculopunctor is shown in Figure 1.

**Figure 1.**
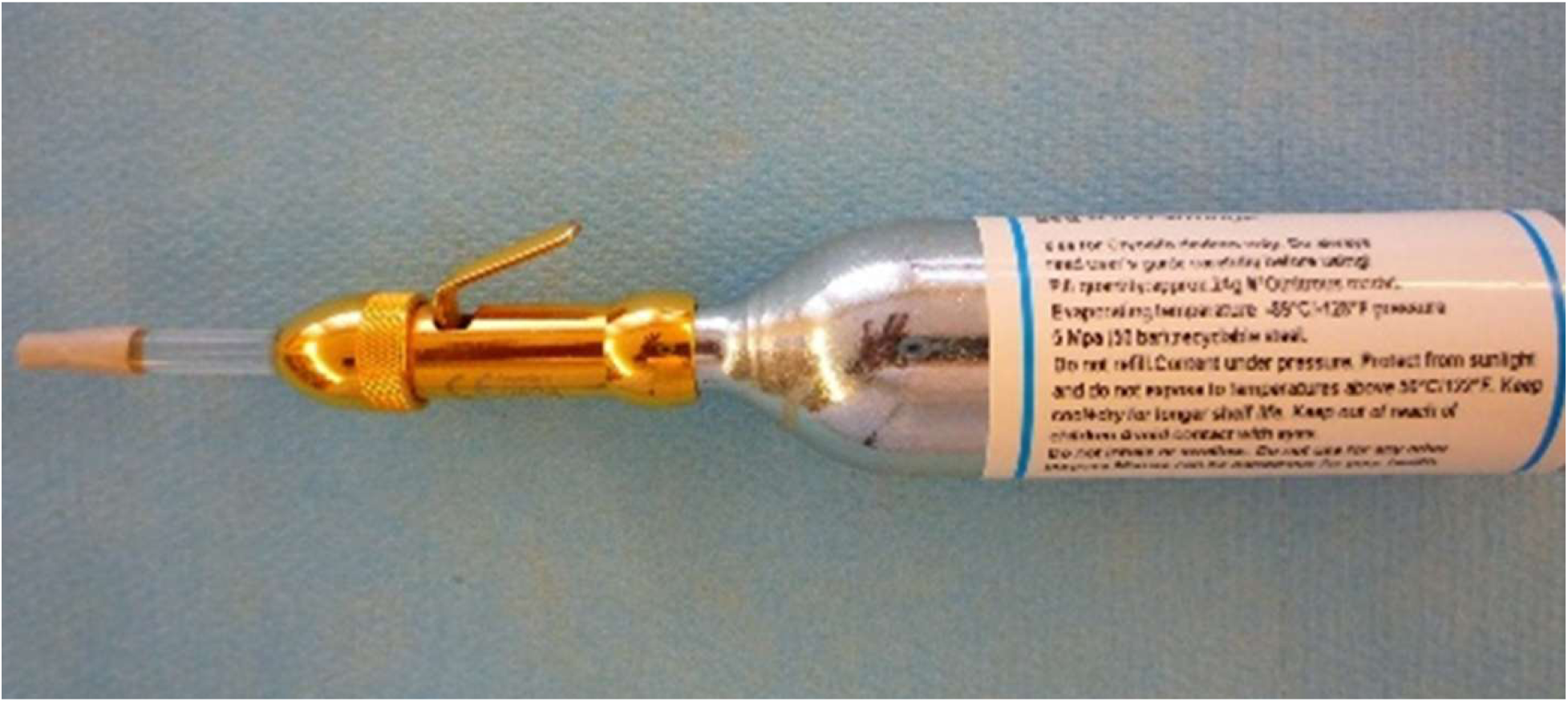
Cryoauriculopunctor composed of one injector connected to a nitrogen canister

### Anesthesia and postoperative analgesia

Prior to transfer to the operating room, an interscalene block (12 ml of bupivacaine 0.5%) using ultrasound was performed on each subject. The surgery was performed under general anesthesia. Except for the trained research staff who performed the auriculotherapy treatment, the subject, the healthcare team (recovery room nurses, surgeons, members of the acute pain team), and research coordinator collecting data were all blinded to the subject’s treatment allocation.

At the time of discharge from the hospital, each subject was given an opioid prescription for five days and instructed to take acetaminophen and ibuprofen for mild to moderate pain and opioids for pain rated as severe (scale: 0-3 =mild pain; 4-6= moderate pain; 7-10= severe pain) and instructed to complete and return a diary on postoperative days 1, 2, 3, 4, and 5. Each subject was called on postoperative 14 days, one month, two months, and three months.

### Data Collection and Outcome Measures

#### Primary outcome

Opioid consumption was collected every day following discharge and is expressed as total opioid consumption from postoperative day 1 to day 5 in oral morphine equivalent (OME in mg). Pain scored using a verbal analogue scale (VAS; 0=no pain to 10=worth possible pain) was also obtained from postoperative days 1 to 3 months. From postoperative day 1 to 5, pain was expressed as the area under the curve.

#### Secondary outcome

Time to discharge from the recovery room, time to discharge from the hospital, pain, and opioid consumption on postoperative day 1, 2, 3, 4, and 5 were recorded. In addition, overall non-narcotic analgesic consumption on postoperative day 1 to day 5, and pain and patient satisfaction related to pain management (0= very dissatisfied to 6= very satisfied) on postoperative day 1, 2, 3, 4, and 5 and 14 days, one month, two months, and three months. Functional recovery was evaluated using the 12-Item Short-Form Health Survey (SF-12) completed prior to surgery and at three months following the surgery. The number of subjects from each group readmitted to the hospital because of pain was also collected.

### Statistical Analysis

Data were analyzed using a modified intend-to-treat analysis. ^(21)^ Power analysis accounted for 10% of subjects not returning their diary as loss of follow up. We estimated that under these conditions, a total of 50 patients would provide 80% power. A non-paired T-test was performed to determine the significance of the difference between the two groups. P < 0.1 was considered significant. Data are expressed as mean ± standard deviation (SD).

## RESULTS

A total of 50 subjects signed an informed consent form. Among them, 10 subjects did not return a completed pain diary to report their pain scores and opioid consumption post-operatively. Another subject was excluded because it was found postoperatively that the subject had a history of chronic opioid use (assigned to the placebo group). Consequently, a total of 39 subjects (20 subjects in the auriculotherapy group and 19 subjects in the placebo group) were included in the final analysis. Figure 2 presents a flow chart of the subjects who signed informed consent. Table 1 presents the demographic characteristics of subjects included in the final analysis for both groups (age, gender, height, weight, body mass index, and racial distribution). The use of auriculotherapy was associated with a significant 35% overall reduction in opioid requirement over the five-day study period compared to the placebo treatment: 62 mg ± 46 OME mg, vs. 96 mg ± 67 OME mg, respectively, (p=0.0307). This reduction in opioid requirement in the treatment group was associated with a significant 16% decrease in pain with movement (21.08 ± 8.62 vs 24. 97 ± 11.29, auriculotherapy vs placebo, respectively; p=0.0827) but no significant difference in pain at rest (15.65 ± 7.48 vs 15.47 ± 9.99, auriculotherapy vs placebo, respectively; p=0.4752). Furthermore, after post-operative day one, 20% of subjects in the active auriculotherapy group versus 11% in the placebo group didn’t use any opioids over the remainder of the five-day study period.

**Table 1.** Demographics of the Auriculotherapy and placebo groups (subjects included in the final analysis)

|  | Auriculotherapy Group<br>(n=20) | Placebo Group<br>(n=19) |
| --- | --- | --- |
| Age (years) | 63.2 ± 9.8 | 63.6 ± 7.6 |
| Gender (M/F) | 9/11 | 10/9 |
| Height (cm) | 169.3 ± 10.3 | 169.1 ± 12.5 |
| Weight (kg) | 89.9 ± 27.1 | 85.0 ± 14.0 |
| BMI (kg/m<sup>2</sup>) | 31.0 ± 6.8 | 29.8 ± 2.6 |
| Race |  |  |
| Asian | 0 | 0 |
| African American | 1 (5%) | 0 |
| Caucasian | 19 (95%) | 19 (100%) |
| Other | 0 | 0 |

Age (mean ± SD), gender (male/female), height (mean ± SD), weight (mean ± SD), Body Mass Index (BMI, mean ± SD) and race distribution (percentages).

**Figure 2.**
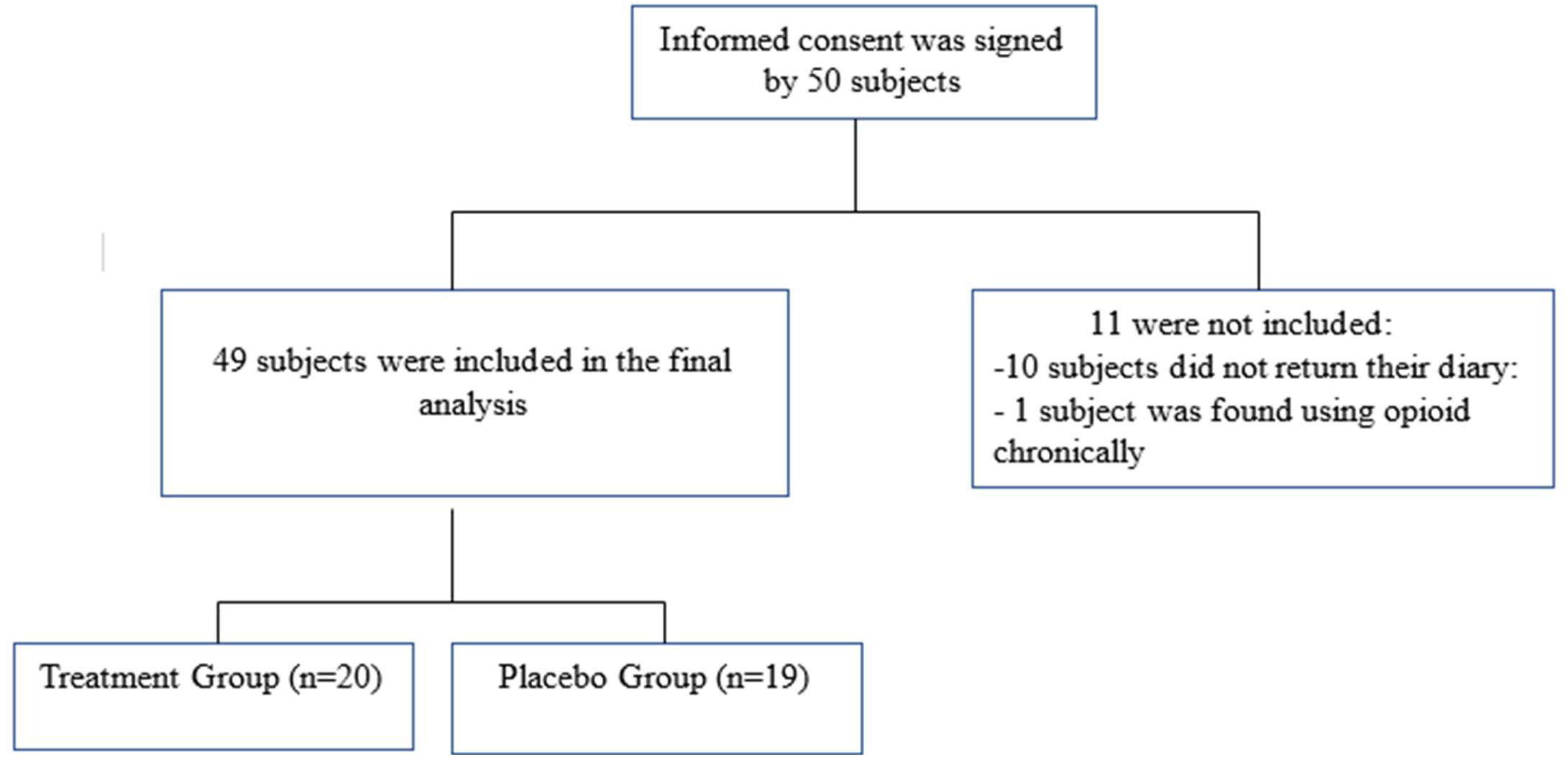
Study flowchart with subjects who were enrolled and included in the final analysis.

Pain at rest was similar in both groups at the time of discharge from the hospital, (0.45 ± 0.76 auriculotherapy vs 0.89 ± 1.91 placebo; p=0.1707) and on the first postoperative day (5.75 ± 3.32 auriculotherapy vs 6.17 ± 3.01 placebo; p=0.3446). Pain at rest and during movement progressively decreased in the subjects receiving an active auriculotherapy treatment over the first four days (3 ± 2.4 changes between day 1 vs. day5 p=0.0085). On day 14, pain during movement was significantly lower in the auriculotherapy group vs the placebo group (4.47 ± 2.12 in vs 5.84 ± 2.39; p=0.0394), suggesting a long-lasting analgesic effect of the auriculotherapy. Figure 3 shows pain ratings at rest on postoperative days 14, 30, 60, and 90. Figure 4 shows pain scores with movement on postoperative days 14, 30, 60, and 90.

**Figure 3.**
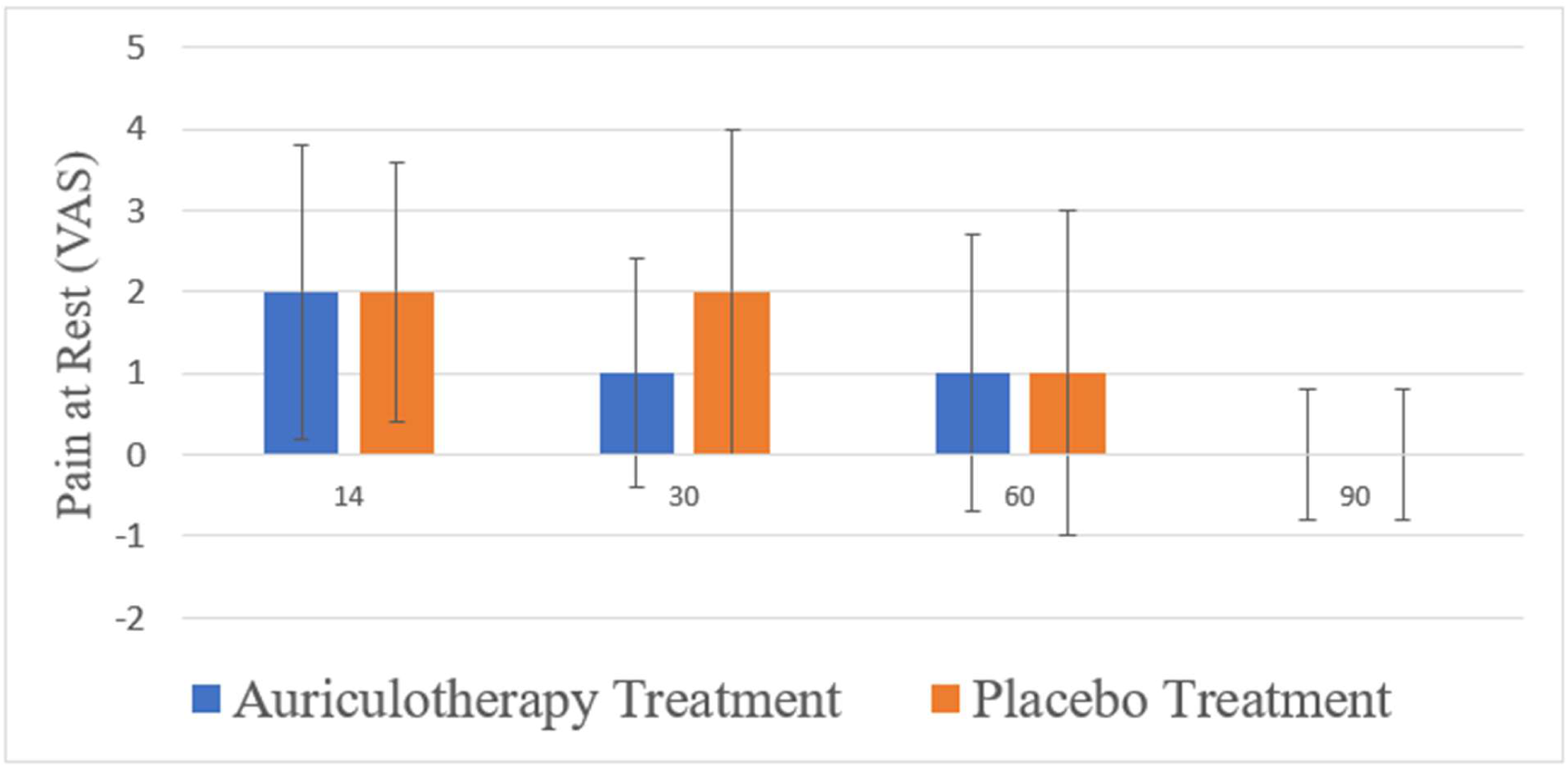
Pain scores on visual analogue scale (VAS) at rest (mean ± SD) on days 14, 30, 60, and 90 after discharge from the hospital

**Figure 4.**
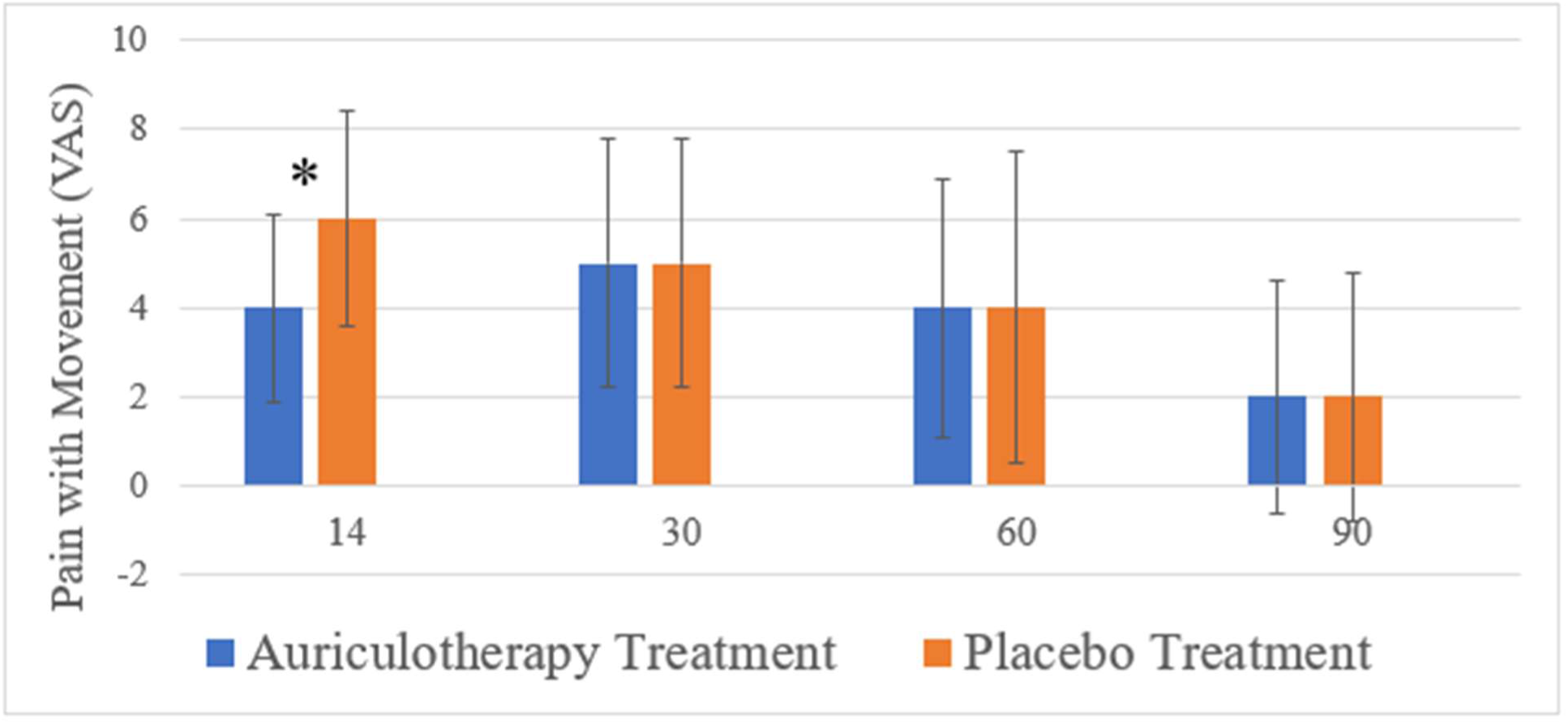
Pain scores on visual analogue scale (VAS) with movement (mean ± SD) on day 14 (p= 0.0394) and days 30, 60, and 90 after discharge from the hospital

No difference between groups was recorded with respect to non-opioid analgesic consumption over the study period (acetaminophen: 4438 ± 4066 in the placebo group vs. 5956 ± 5882 in the auriculotherapy group, p=0.1786; ibuprofen 785 ± 1577 in the placebo group vs. 1370 ± 2386 in the auriculotherapy group, p=0.1858).

No difference between groups was recorded with respect to non-opioid analgesic consumption over the first 5 postoperative days (acetaminophen: 4438 ± 4066 in the placebo group vs. 5956 ± 5882 in the auriculotherapy group, p=0.1786; ibuprofen 785 ± 1577 in the placebo group vs. 1370 ± 2386 in the auriculotherapy group, p=0.1858). No difference between groups was recorded with respect to either discharge time from the recovery room (67.2 ± 34.1 min vs. 59.5 ± 25.7 min, auriculotherapy vs. placebo group, respectively; p=0.2220) or time to discharge from the hospital (148.0 ± 47.4 min vs. 140.1 ± 32.1 min, auriculotherapy vs. placebo group, respectively; p=0.2856).

Overall patient satisfaction with pain treatment was found to be similar in both the auriculotherapy and placebo groups, except on day 90 when subjects in the auriculotherapy group reported being more satisfied than those in the placebo group. Table 2 reports the patient satisfaction scores throughout the study period. Lastly, functional recovery assessed with the SF12 was found to be similar in both the active and placebo groups over the study period and no subject in either group was reported to be readmitted to the hospital due to pain during the study period.

**Table 2.** Overall patient satisfaction with pain management in the auriculotherapy and placebo group

| Postoperative period | Auriculotherapy Group<br>(n=20) | Placebo Group<br>(n=19) | p-value |
| --- | --- | --- | --- |
| Day 1 | 4.1 ± 1.4 | 4.2 ± 1.8 | 0.3749 |
| Day 2 | 4.6 ± 1.1 | 4.7 ± 1.2 | 0.3862 |
| Day 3 | 4.8 ± 1.0 | 5.1 ± 0.8 | 0.1735 |
| Day 4 | 4.6 ± 1.3 | 4.7 ± 1.5 | 0.4304 |
| Day 5 | 4.8 ± 1.1 | 4.8 ± 1.5 | 0.4902 |
| Day 14 | 5.6 ± 0.8 | 5.6 ± 0.8 | 0.4371 |
| Day 30 | 5.7 ± 0.5 | 5.4 ± 1.2 | 0.1950 |
| Day 60 | 5.3 ± 1.5 | 5.2 ± 1.6 | 0.4124 |
| Day 90 | 5.9 ± 0.3 | 4.9 ± 1.9 | 0.0268* |

\* indicates statistically significant, with $p < 0.1$

## DISCUSSION

This randomized, placebo-controlled study demonstrated that the use of auriculotherapy based on Dr. David Alimi’s cartography ^(20)^ allowed for a 35% reduction in opioid requirement in opioid-naïve subjects following rotator cuff surgery with a preoperative interscalene block. Since the interscalene block lasts up to 24 hours, it is not surprising that no difference in pain between the two groups was observed on postoperative day 1. The progressive decrease in pain observed in the auriculotherapy group demonstrated that the optimum effect of auriculotherapy was reached on postoperative day 4. Although, this may represent an argument in favor of performing the auriculotherapy treatment prior to surgery, performing auriculotherapy a few days prior to surgery is not practical because many patients live far away from the hospital.

Dr. Alimi’s cartography ^(20)^ was initially presented and accepted by the World Federation of Chinese Medicine and presented at the World Congress of Chinese Medicine on September 3, 2011. It is based on the use of a segmentogram using the corpus callosum as its center of symmetry and 189 points on the lateral aspect of the ear and 89 points on the internal aspect of the ear. This nomenclature is side-specific, meaning that the representation of the right ear includes the point of the liver, whereas the left side includes the point of the pancreas. The proposed cartography has been validated by MRI studies, including studies validating the point of the thumb ^(22,23)^ and the knee. ^(24)^

According to the French school of auriculotherapy led by Alimi, the auriculotherapy treatment is based on the physiopathology of the conditions being treated, meaning that in the case of this study, the points chosen to treat postoperative pain included the master point for the mesoderm (tissue of origin for the musculoskeletal system), the site of the surgery, and points involved in the pain pathway and inflammation. ^(25)^ Such an approach is different than the one proposed by Chinese acupuncture, which is based on the use of ear points according to meridians and energy (Yan and Yin). ^(26)^

Each ear point corresponds structurally to a Merkel disk, ^(27)^ which includes a terminal branch of one of the cranial nerves innervating the ear. Each of these points exists in two states: “physiologic” (characterized by a given difference of potential and the absence of pain) or “pathologic” (characterized by an abnormal difference of potential and the presence of pain) on contact. The status of each point can be established using a galvanometer or by assessing the degree of pain. The terminal nerve branch present at the level of each ear point transmits impulses to the cranial nerve they originate from (the vagal, trigeminal, facial, and glossopharyngeal nerves, or the superficial cervical plexus) ^(28-29)^ located at the level of the spine and brainstem. These nuclei interact with ascendant and descendent pathways to “communicate” the central nervous system and the periphery. ^(30)^ and interact with a complex network of central nervous system structures including the amygdala, thalamus, and nucleus of tractus solitary. ^(31-33)^ The destruction of the nerve terminal of a Merkel disk by an acupuncture needle stimulates the brain to regenerate the disk in its “physiological” according to the third law of D’Otto Khaler, ^(34)^ meaning in this case in its “physiologic” state and also re-establish the peripheral status of the organ the point represents.

The cryoauriculopunctor we used in this study was developed by Dr. Alimi. ^(35)^ It allows for the use of gas instead of needles to interact with the auricular treatment points. Prior studies have shown that in different models, the use of gas produces the same therapeutic response as needle stimulation (Alimi, personal communication). This presents several advantages. First, the use of gas instead of a needle is often better accepted by a patient, especially when they are phobic of needles, and second, it allows for the performance of a true placebo-controlled trial. Thus, in the case of a placebo treatment, the same point is treated using an empty gas canister, resulting in no gas being dispensed. This process gives the patient the impression that a treatment is being administered. When real needles are used and a randomized placebo-controlled trial is performed, the placebo treatment points would consist of placing the needles in “non-active areas of the ear,”^(17)^ with the assumption that the points chosen are “non-active.” This is a very arbitrary assumption that may explain the fact that a number of randomized, placebo-controlled acupuncture trials are unable to demonstrate a difference between the active and placebo treatments. ^(9)^

## CONCLUSION

Cryogenic auriculotherapy, a neurophysiological-based treatment using Dr. Alimi’s cartography, was employed as a single treatment just after rotator cuff surgery, during which patients also received an interscalene peripheral nerve block. Using a randomized, placebo-controlled design in this patient population resulted in a reduction in postoperative opioid requirement and reduction in pain with movement for at least 14 days after surgery. Additional studies are needed to further characterize this effect and determine the optimal number and timing of cryogenic auriculotherapy treatments to minimize postoperative pain and opioid use.

## Data Availability

All data produced in the present study are available upon reasonable request to the authors.

## ACKNOWLEDGEMENTS

We want to thank Mrs. Christine Heiner for editing this manuscript.

## AUTHOR CONFIRMATION STATEMENT

All named authors meet the International Committee of Medical Journal Editors (ICMJE) criteria for authorship for this article, take responsibility for the integrity of the work as a whole, and have given their approval for this version to be published.

## DATA AVAILABILITY

The datasets generated during and/or analyzed during the current study are available from the corresponding author upon reasonable request.

